# A Detector Block-Pairwise Dead Time Correction Method for Improved Quantitation with a Dedicated BrainPET Scanner

**DOI:** 10.1101/2022.09.12.22279839

**Authors:** Ahlam Said Mohamad Issa, Jürgen Scheins, Lutz Tellmann, Alejandro Lopez-Montes, Joaquin L Herraiz, Cláudia Régio Brambilla, Hans Herzog, Irene Neuner, N. Jon Shah, Christoph Lerche

**Affiliations:** Institute of Neuroscience and Medicine 4, INM-4, Forschungszentrum Jülich, Jülich, Germany; JARA - BRAIN - Translational Medicine, Aachen, Germany; Department of Neurology, RWTH Aachen University, Aachen, Germany; Nuclear Physics Group and IPARCOS, University: Complutense University of Madrid, Madrid, Spain; Department of Psychiatry, Psychotherapy and Psychosomatics, RWTH Aachen University, Aachen, Germany; Institute of Neuroscience and Medicine 11, INM-11, JARA, Forschungszentrum Jülich, Germany

**Keywords:** dead time, PET, block-pairwise, MR/PET systems, delayed random coincidence, time-activity curves

## Abstract

Dead time correction (DTC) is an important factor in ensuring accurate quantification in PET measurements. This is currently often achieved using a global DTC method, i.e., an average DTC factor is computed. For PET scanners designed to image dedicated organs, e.g., those used in brain imaging or positron emission mammography (PEM), a substantial amount of the administered radioactivity is located outside of the PET field-of-view (FOV). This activity contributes to the dead time (DT) of the scintillation detectors. Moreover, the count rates of the individual scintillation detectors are potentially very inhomogeneous due to the specific irradiation of each detector, especially for combined MR/PET systems, where radiation shields cannot be applied. We have developed a block-pairwise DTC method for our Siemens 3T MR BrainPET insert by extending a previously published method that uses the delayed random coincidence count rate to estimate the DT in the individual scans and planes (i.e., scintillation pixel rings). The method was validated in decay experiments using phantoms with a homogenous activity concentration and with and without out-of-FOV activity. Based on a three-compartment phantom, we compared the accuracy and noise properties of the block-pairwise DTC and the global DTC method. While the currently used global DTC led to a substantial positive bias in regions with high activity, the block-pairwise DTC resulted in substantially less bias. The noise level for the block-pairwise DTC was comparable to the global DTC and image reconstructions without any DTC. Finally, we tested the block-pairwise DTC with a data set obtained from volunteer measurements using the mGlu5R (metabotropic glutamate receptor subtype 5) antagonist [^11^C]-ABP688, when comparing the time-activity curves (TACs) obtained with the global DTC with the block-pairwise DTC, relative differences in the anterior cingulate cortex (ACC) and the cerebellum of up to 25% were observed during the first 30 minutes of these measurements.

## 1. Introduction

One of the most relevant sources of errors in PET imaging is count losses caused by the DT of the scintillation detectors and the data acquisition system (DAQ) (Meikle and Badawi 2005, Leo 2012, Uribe and Celler 2022, Cherry and Dahlbom 2006, Usman and Patil 2018, Scheins *et al* 2018). For quantitative PET imaging, the accurate correction of these losses is indispensable, in addition to other data corrections such as attenuation correction, scatter correction and random coincidence correction. The most relevant components in a PET system that suffer from DT are the scintillation detector, the digitization electronics, and the coincidence electronics (Knoll 2010). DT losses affect the reconstruction results of quantitative images by causing an underestimation of the pixel counts (Mazoyer *et al* 1985, Freedman *et al* 1992). Furthermore, the DT losses increase at high count rates and vary during the scan time as the radiotracer distributes and decays. This is particularly significant for dynamic studies that require the exact quantification of the injection bolus and the use of tracers with a short half-life (Hoffman *et al* 1983, Zaidi 2006). Insufficient DTC will result in distortion of the measured TACs and quantities derived from the TAC, such as the distribution volume and the binding potential (BP_ND_). Finally, inappropriate corrections invalidate the assumption of the Poisson nature of the detected events and may lead to reconstruction and quantification biases when using maximum likelihood (ML) based reconstructions (Toga *et al* 2002, Morris *et al* 2004, Brambilla *et al* 2021.). The Siemens 3T MR-BrainPET insert at Forschungszentrum Jülich is one of several prototypes of dedicated hybrid MRI/PET systems worldwide. It is built from a commercial 3T MRI and an MR-compatible BrainPET insert to enable the simultaneous acquisition of structural and molecular images as previously shown (Herzog *et al* 2011, Shah *et al* 2013, Herzog 2012). The system applies a global DTC method. This method is based on a count rate-dependent correction factor, which is obtained by averaging over all scintillation detector blocks. For this purpose, the constant fraction discriminator (CFD) counts of each block are summed over all scintillation detectors to estimate the global, non-validated single counts and the overall single count losses caused by DT as previously shown (Weirich et *al* 2012, Caldeira et *al* 2018, Weirich et *al* 2013, Weirich et *al* 2013). The Siemens 3T MR-BrainPET insert consists of 192 scintillation detector blocks arranged in 32 cassettes. Each cassette has six block detectors, thus forming six block detector rings. Figure 1 shows the unqualified, uncorrected single count rates for eight different cassettes in one sector (left) and the six different rings (right) from a volunteer measurement with [^11^C]-ABP688 (Régio Brambilla et *al* 2020). It can be seen that the uncorrected count rates are significantly different for the individual cassettes and rings. The highest single count rates were found in those blocks close to the radioactivity sources, especially in the first ring of the PET insert. The large differences observed in these count rates affect the accuracy of the DTC factor of the global DTC method, as DT losses depend non-linearly on the count rates and the differences are not correctly accounted for when using averaged DTC factors. Furthermore, the physical position of each block is ignored. This results in a significant loss in the quantitation accuracy of the DTC. Thus, differences in the temporally and spatially varying irradiation when imaging living subjects will lead to quantification errors (Mazoyer *et al* 1985). Apart from the non-uniform activity distribution inside the BrainPET FOV, the DT of individual block detectors is also affected by the radiation from the areas of the body outside the FOV. This further increases the differences in the single count rates seen by the individual detector blocks and makes the DT losses dependent on the position of the detector block. Therefore, correcting the DT losses for each individual block should provide more accurate results for quantitative imaging.

**Figure 1.**
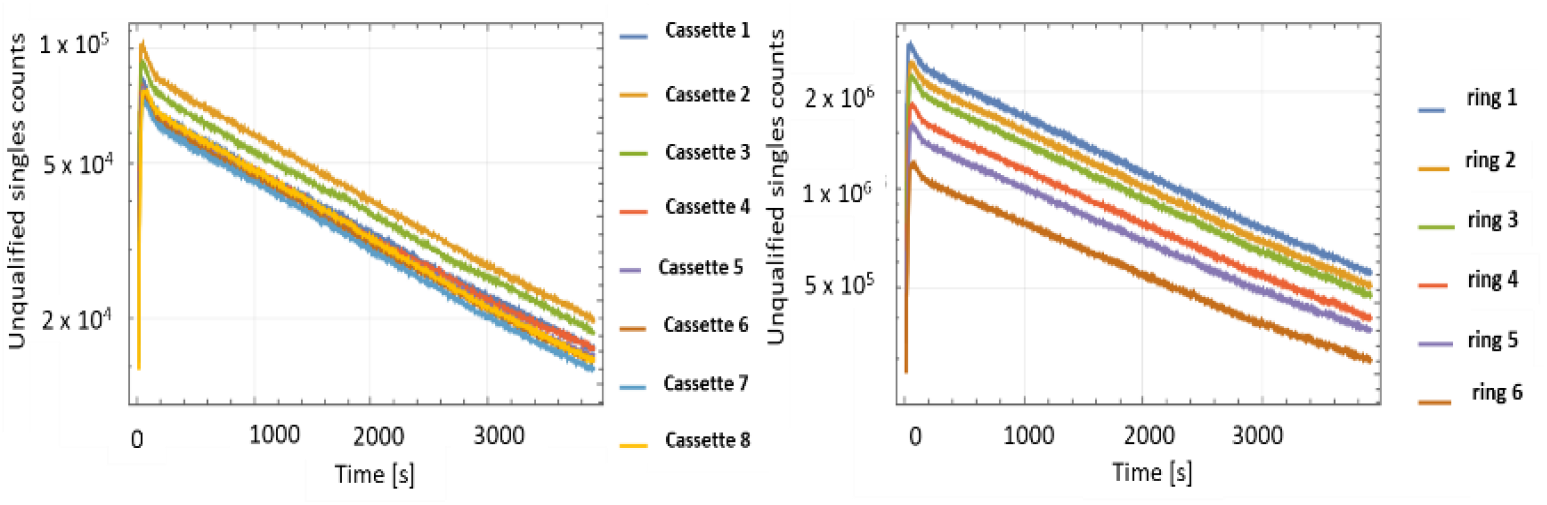
The unqualified single counts for the eight detector cassettes of one sector on the left side and for the individual block rings on the right side. Measurements were taken from an [^11^C]-ABP688 volunteer measurement using the Siemens 3T MR-BrainPET insert.

Figure 2 shows the reconstructed activity concentration in Bq/cm^3^ for a phantom measurement over several half-lives of the radioactive tracer nuclide. The phantom was filled with an ^18^F solution and placed inside of the PET FOV.

**Figure 2.**
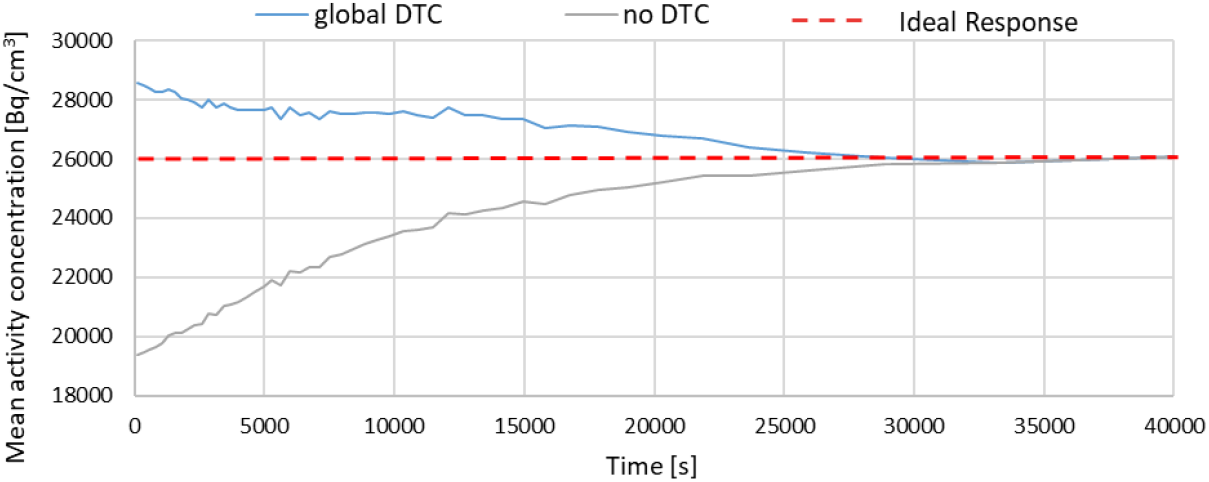
Reconstructed activity concentration in Bq/cm^3^ vs time in seconds for a phantom measurement (^18^F) with an ROI on the back of the FOV and with out-of-FOV activity.

An additional ^18^F phantom with the same level of activity was placed outside of the FOV. A region of interest (ROI) was drawn in the reconstructed image of the phantom - located on the back of the phantom. One can see that the global DTC method produces a significant overcorrection of up to 10% compared to the expected reconstructed activity concentration (dashed red line). For comparison, the reconstructed activity concentration without DTC is also shown.

The main motivation of this work was to improve the accuracy of the DTC method by implementing a block-pairwise DTC method where the DT is corrected for each pair of scintillation blocks individually. The presented method is based on the estimation of the DTC factor as a nonlinear function derived from the delayed random coincidence rate for individual detector rings (Yamamoto *et al* 1986). The method was adapted for estimating the DT losses for the 3T MR-BrainPET insert for individual scintillation block pairs instead of individual rings. Further, the method was also modified by estimating the DT parameter by fitting the observed count rates according to a theoretical model as a function of activity over time. Finally, we found that randomly-detected triple coincidences (Lage *et al* 2015, Cal-González *et al* 2014, Pál and Pázsit 2012) should be considered to improve the accuracy of the method. In this work, the precision (statistical noise) and the accuracy of this modified method are compared with the global DTC method currently used on the Siemens 3T MR-BrainPET insert.

## 2. Materials

### 2.1 Dead time correction method

The large differences in block count rates observed in typical volunteer measurements (see figure 1) are the main reasons to implement a DTC method on a block-wise level. For the implementation, both standard mathematical models for DT, i.e., the paralyzable and non-paralyzable models (Müller 1973, Bailey *et al* 2005, Ensslin 1991) were tested. As will be shown, the count rates observed during typical PET examinations with the Siemens 3T MR-BrainPET insert are not high enough to lead to substantial differences between the models. Due to the absence of differences, the block-wise DTC method based on the non-paralyzable (NP) model was implemented, and only the derivation of the DTC factor for the NP model is described in this work. However, the derivation of the DTC factor for the paralyzable model is analog. As shown in (Yamamoto *et al* 1986), the expected delayed random coincidence rate without DT losses can be obtained by extrapolation of the observed delayed coincidences. This allows the computation of a DTC factor for the observed prompts coincidence rate (Yamamoto *et al* 1986). The observed delayed random coincidences rate (*R*_*ob*_), measured with the delayed window technique, (Markiewicz *et al* 2018, Markiewicz *et al* 2016) is affected by the DT as follows:

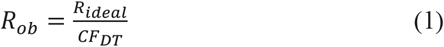

where *R*_*ideal*_ is the delayed random coincidences rate without DT losses for any pair of scintillation blocks, and the *CF*_*DT*_ is the DTC factor. Equation (1) allows computation of the correction factor *CF*_*DT*_, if *R*_*ideal*_ and *R*_*ob*_ are known during every second of the measurement. *P*_*corr*_ are the corrected prompt coincidence count rates for each pair of scintillation blocks, and they are given by the sum of *R*_*ideal*_ and the corrected true coincidence count rates, *T*_*corr*_. *T*_*corr*_ are given by:

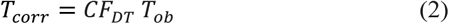

where *T*_*ob*_ denotes the observed true coincidence count rates. Equally, the corrected prompt coincidence count rates, *P*_*corr*_, can be computed by:

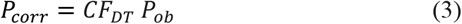

where *P*_*ob*_ are the observed prompt coincidence count rates. Expression (4) gives the expected count rates in the case of NP DT:

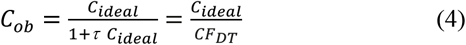

where *C*_*ob*_ is the observed count rate. *C*_*ideal*_ is the count rate without DT losses for any pair of scintillation blocks and *τ* is the DT. For a decaying activity, the ideally expected delayed random count rates as a function of the time, *t*, can be parametrized as follows:

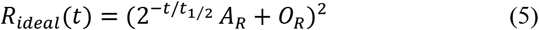

where *A*_*R*_ is the random count rate due to the activity in the phantom at *t*=0, *t*_1/2_ is the half-life of the radioactive tracer, and *O*_*R*_ is a constant offset term which accounts for random counts caused by background radiation and the natural, intrinsic ^176^Lu activity (Alva-Sánchez *et al* 2018). One important effect which is considered in this work, in addition to the method presented in (Yamamoto *et al* 1986), is the correction of triple coincidences, i.e., when at least three gamma photons are detected within the coincidence timing window. Triple coincidences are not uncommon and may be caused by inter-detector scatter and random events (Lage *et al* 2015, Cal-González *et al* 2014, Pál and Pázsit 2012). In the Siemens 3T MR-BrainPET insert, triple coincidences are found as two double coincidences in the list-mode data and are found both in the prompt and delayed coincidences lists. Thus, they result in count rate overestimation, and a corresponding triples correction should be applied to correct this effect, especially as the coincidence window of the BrainPET insert is rather large - 12 ns. Due to their much lower probability, correction for multiple coincidences was not explicitly considered by the manufacturer. The overestimation of delayed random counts caused by triples can be approximated by the empirical model:

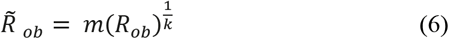

where 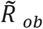 are the observed counts with overestimation due to triples and *m* and *k* are free fit parameters. The expected counts of delayed random coincidences, *R*_*ob*_, can now be expressed by combining equations (4), (5), and (6):

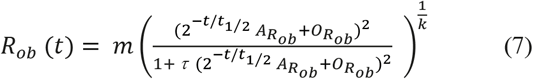

where 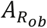 is the count rate for the observed delayed coincidence and corresponds to the tracer activity in the phantom at *t*=0. 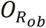 is the corresponding pedestal accounting for natural background radiation. The DTC factor, *CF*_*DT*_, can now be obtained in two steps. First, equation (7) is solved for *t* resulting in equation (8):

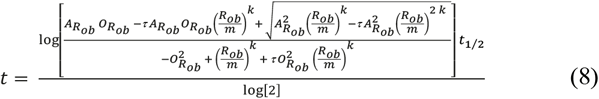

Then, equation (8) is inserted into (5), thus yielding equation (9):

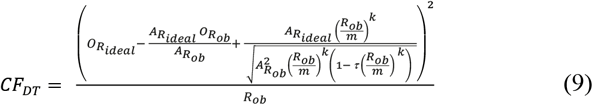

where 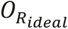 is introduced as the independent pedestal for ideal delayed random coincidences and 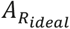 is the count rate at *t*=0 for ideal delayed random coincidences. Equation (9) gives the correction factor, *CF*_*DT*_, for one single pair of scintillation blocks. As the Siemens 3T MR-BrainPET insert has six rings, 32 cassettes, and coincidences with only 19 of 32 cassettes are used, the *CF*_*DT*_ for 6^2^ × 32 × 19 × 1/2 = 10944 must be determined and applied. Thus, the *CF*_*DT*_ represents correction factors for block pairs and not for single blocks. All variables in equation (9), except the observed delayed random count rate *R*_*ob*_, are used as free fit parameters. For the calibration, the time-varying delayed count rates for all 10944 block-pairs were fitted with equation (9), and the best fit parameters were stored. For correction during measurements, the 10944 individual count rates *R*_*ob*_ were used together with the corresponding set of best fit parameters, and equation (9) was used to compute the DTC factor for each pair, which was then subsequently used to correct the prompt count rate for the corresponding block-pair according to equation (3).

### 2.2 Data acquisition

All measurements were performed in the Siemens 3T MR-BrainPET insert. The data were recorded in list-mode format and gathered into sinograms. Random coincidences were estimated by the delayed window technique (Markiewicz *et al* 2018, Markiewicz *et al* 2016). The energy acceptance window was set to 420 - 600 keV, while the coincidence time window was 12 ns. The BrainPET insert uses avalanche photo-diodes (APDs), and scintillation event processing is done on the Quicksilver platform (Newport *et al* 2006, Hu *et al* 2011). For temperature stabilization, the detector modules are supplied with cooled air. LSO is used as the scintillator. The LSO crystals have a pixel pitch of 2.5 mm x 2.5 mm, and their length is 20 mm. The crystals are coupled by a light guide to a 3 × 3 array of Hamamatsu S8664–55 APDs, each with a sensitive area of 5×5 mm^2^. The PET scanner consists of 192 detector blocks arranged in 32 cassettes (heads), each cassette bearing six detector blocks. Each detector block, with its 12×12 scintillation pixels, the 3×3 APDs, and the attached analog readout electronics, acts as one position-sensitive scintillation detector with an individual count rate and DT loss. The BrainPET insert has an axial FOV of 19.2 cm and a transversal FOV of 31.4 cm (Weirich et al 2012, Caldeira et al 2018, Weirich et al 2013, Weirich et al 2013).

### 2.3 Measurements

Phantom measurements with decaying activity and with a known half-life were used to calibrate, validate, and evaluate the presented DTC method. During the phantom measurements, prompt and delayed count rates were registered for every second during approximately 7 half-lives (≈ 20 hours). For each block-pair, the DTC factor is computed and applied to the prompts. For these decay experiments, cylindrical, homogeneous phantoms were used. For the first measurement (I), a homogeneous cylinder with a 14 cm inner diameter and 23.6 cm length (volume: ≈ 3633 ml) was filled with high activity (257.4 MBq) ^18^F diluted in water. The phantom was transversally centered in the FOV within the MR transmit/receive head coil. For the second measurement (II), a homogeneous cylinder with an inner diameter of 14 cm and a length of 23.6 cm (volume: ≈ 3633 ml) was filled with high activity (141.8 MBq) ^18^F diluted in water. The phantom was transversally centered in the FOV. The phantom was axially centered to the extent that the dimensions of the MR transmit/receive head coil allowed. In addition, a second homogeneous cylinder phantom with an inner diameter of 20 cm and length of 19 cm (volume ≈ 5969 ml) was filled with high activity (283 MBq) ^18^F diluted in water and placed outside of the FOV. Thus, the total activity was divided into two parts. Thirty percent was placed inside of the FOV, and the remaining (70%) was placed outside of the FOV to mimic uptake in the subject’s body, which contributes significantly to the detector DT. For the performance evaluation of the presented method, a third measurement (III) using an adapted three-compartment phantom (NEMA PET phantom TM (NU 2-994)) filled with ^18^F diluted in water was performed. The inner diameter of the background compartment was 20 cm, and the length was 23 cm (volume ≈ 4180 ml). The three inserted compartments were 5 cm in diameter and 20 cm in length (volume ≈260 ml). One of them was filled with an activity concentration six times higher than the background compartment (hot compartment), and one was filled with an activity concentration double times higher than the background (warm compartment). One insert was made of Teflon (cold compartment). The ^18^F activity in the background compartment was 113.3 MBq. Data were acquired for 11 hours, and the phantom was centered inside of the FOV. In addition, a second homogeneous cylinder phantom (dimensions as described above) was filled with 82.6 MBq of ^18^F activity and was placed outside of the FOV.

The compartment to background ratios for the hot and warm compartments were 6:1 and 2:1, as these values represent typical values for the tumor to background ratios and cortex region to cerebellum ratio, respectively. We also studied the impact of the out-of-FOV activity on the calibration factor and its homogeneity for both methods (global DTC and block-pairwise DTC) by evaluating the cross-calibration factor in several ROIs in calibration scans taken with and without out-of-FOV activity. The fourth measurement was a calibration measurement (IV), and it was divided into two sessions. In the first one, a homogeneous cylinder with an inner diameter of 14 cm and a length of 23.6 cm (volume: ≈ 3633 ml) was filled with high activity (≈ 64 MBq) ^18^F diluted in water. The phantom was transversally centered in the FOV within the MR transmit/receive head coil. In addition, a second homogeneous cylinder phantom with an inner diameter of 20 cm and a length of 19 cm (volume ≈ 5969 ml) was filled with high activity (≈ 126 MBq) ^18^F diluted in water and placed outside of the FOV. The scan time was 30 minutes. The second session used the phantom in the FOV without any activity outside of FOV (after 30 minutes of the activity decay). The decay of the tracer between both measurements was corrected. The aim of the measurement was to quantify the effects of the activity outside of FOV (i.e., the effect of a patient’s whole body on brain imaging) on the calibration factor obtained from the cross-calibration of the system using a well counter. The volunteer’s TAC was obtained with [^11^C]-ABP688 in a PET acquisition of 65 minutes in duration, using a bolus-infusion scheme and a total administered activity of 437 MBq (Régio Brambilla *et al* 2020). Table 1 summarizes all the measurement features.

**Table 1.**
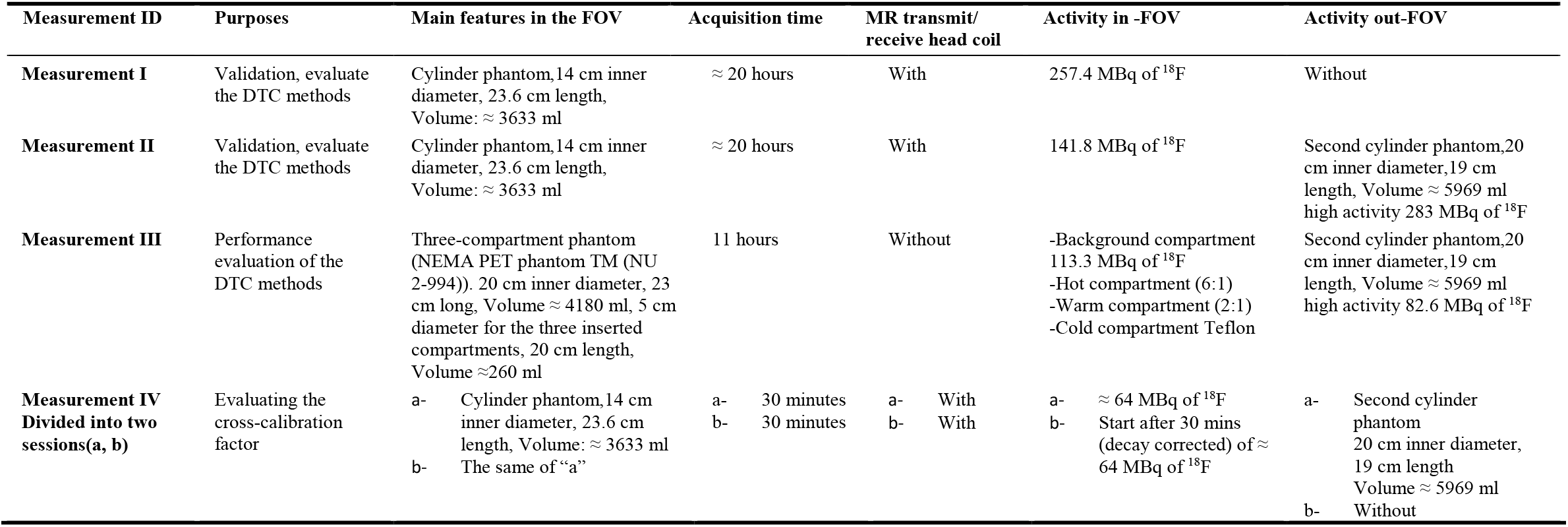
Measurements features of the DTC method validation and calibration.

| Measurement ID | Purposes | Main features in the FOV | Acquisition time | MR transmit/<br>receive head coil | Activity in -FOV | Activity out-FOV |
| --- | --- | --- | --- | --- | --- | --- |
| **Measurement I** | Validation, evaluate the DTC methods | Cylinder phantom, 14 cm inner diameter, 23.6 cm length, Volume: $\approx 3633$ ml | $\approx 20$ hours | With | 257.4 MBq of $^{18}\text{F}$ | Without |
| **Measurement II** | Validation, evaluate the DTC methods | Cylinder phantom, 14 cm inner diameter, 23.6 cm length, Volume: $\approx 3633$ ml | $\approx 20$ hours | With | 141.8 MBq of $^{18}\text{F}$ | Second cylinder phantom, 20 cm inner diameter, 19 cm length, Volume $\approx 5969$ ml high activity 283 MBq of $^{18}\text{F}$ |
| **Measurement III** | Performance evaluation of the DTC methods | Three-compartment phantom (NEMA PET phantom TM (NU 2-994)). 20 cm inner diameter, 23 cm long, Volume $\approx 4180$ ml, 5 cm diameter for the three inserted compartments, 20 cm length, Volume $\approx 260$ ml | 11 hours | Without | -Background compartment 113.3 MBq of $^{18}\text{F}$<br>-Hot compartment (6:1)<br>-Warm compartment (2:1)<br>-Cold compartment Teflon | Second cylinder phantom, 20 cm inner diameter, 19 cm length, Volume $\approx 5969$ ml high activity 82.6 MBq of $^{18}\text{F}$ |
| **Measurement IV**<br>**Divided into two sessions(a, b)** | Evaluating the cross-calibration factor | a- Cylinder phantom, 14 cm inner diameter, 23.6 cm length, Volume: $\approx 3633$ ml<br>b- The same of "a" | a- 30 minutes<br>b- 30 minutes | a- With<br>b- With | a- $\approx 64$ MBq of $^{18}\text{F}$<br>b- Start after 30 mins (decay corrected) of $\approx 64$ MBq of $^{18}\text{F}$ | a- Second cylinder phantom 20 cm inner diameter, 19 cm length, Volume $\approx 5969$ ml<br>b- Without |

### 2.4 Data Analysis

For the validation of the method, the corrected true coincidence rates of all 10944 block-pairs were summed, and the resulting scanner-wide true coincidence rate was fitted to a simple decaying exponential with the ^18^F half-life to determine the relative fit residuals. For validation of the appropriateness of the empirical model for over counting due to triples, all prompt coincidences as a function of the pure double coincidences were fit to equation (6). For the performance evaluation, images were reconstructed with the 3D OP-OSEM algorithm (Mourik *et al* 2010, van Velden *et al* 2008). The image volume matrix was 256 × 256 × 153 pixels with two subsets, 32 iterations, and an isotropic voxel size of 1.25 mm^3^. In addition to the DTC, the data sets were corrected for decay, randoms, attenuation, and scatter (Kops *et al* 2014). The corresponding phantom measurements were reconstructed using global DTC, without any DTC, and with the proposed block-pairwise DTC method. No further data processing was conducted for the phantom measurements. Constant true coincidence count rate framing schemes were used in all reconstructions to minimize reconstruction bias at low counts (Brambilla *et al* 2021, Régio Brambilla *et al* 2022). In the third measurement (III), five circular ROIs were specified in each compartment to enable the evaluation of accuracy and noise level. The same dimensions were used for all the ROIs: x ≈ 44, y ≈ 44, and z ≈ 8 mm. The ROIs were aligned with the compartment’s centric axis (again for all 48 time frames). The ROIs were placed in the center, front, and the back of the phantom (see figure 6). The two ROIs that were not centered in the axial direction were 1cm and 2 cm away from the edges, respectively. For measurement IV (see Table 2), the following ROIs were used. Three circular ROIs were created in the center, front, and the back of the phantom image (ROI surfaces at 1 cm from the edges). The dimensions of the ROIs were x ≈ 81, y ≈ 80, and z ≈ 8 mm, and they were aligned at the scanner axis for all image frames of 30 minutes in length. The ROIs at the front and the back of the phantom have the same distance from the isocenter. The ROI analysis was performed with AMIDE (Amide’s a Medical Imaging Data Examiner software) (Loening and Gambhir 2003). The values used for the ROI analysis are the averaged activity concentration for each frame and the coefficient of variation (COV). The COV was calculated by dividing the standard deviation by the mean of the activity for each midpoint of the period of time (in seconds). The analysis of the [^11^C]-ABP688 PET images was done with PMOD (version 4.103, PMOD Technologies, Zurich, Switzerland), using the PNEURO package. A 3D Gaussian post-reconstruction filter (2.5 mm) was applied. The PET data set was corrected for motion, normalized, and matched to the simultaneously acquired MR T1 MPARGE image. The ROIs were drawn using the T1 MPARGE images as an anatomical reference. Three exemplary regions of the human brain were chosen for the analysis based on their relevance: cerebellum gray matter, temporal posterior lobes, and the ACC. TACs and the non-displaceable BP_ND_ were also analyzed. The BP_ND_ was computed by dividing the mean activity in the target region by the mean activity concentration in the reference region i.e., the cerebellum gray matter, and subtracting the value of 1.0. A detailed description of the volunteer measurements with [^11^C]ABP688 can be found in (Brambilla *et al* 2021, Toga *et al* 2002, Rajkumar *et al* 2021).

**Table 2.** The relative differences when comparing calibration factors with and without out of FOV activity for three relevant ROIs and the global DTC and the block-pairwise DTC method.

| DTC method | Front side of phantom | Center of phantom | Back side of phantom |
| --- | --- | --- | --- |
| global DTC | -4.32% | -1.50% | 0.56% |
| block-pairwise DTC | -2.10% | 1.90% | 0.79% |

**Figure 3.**
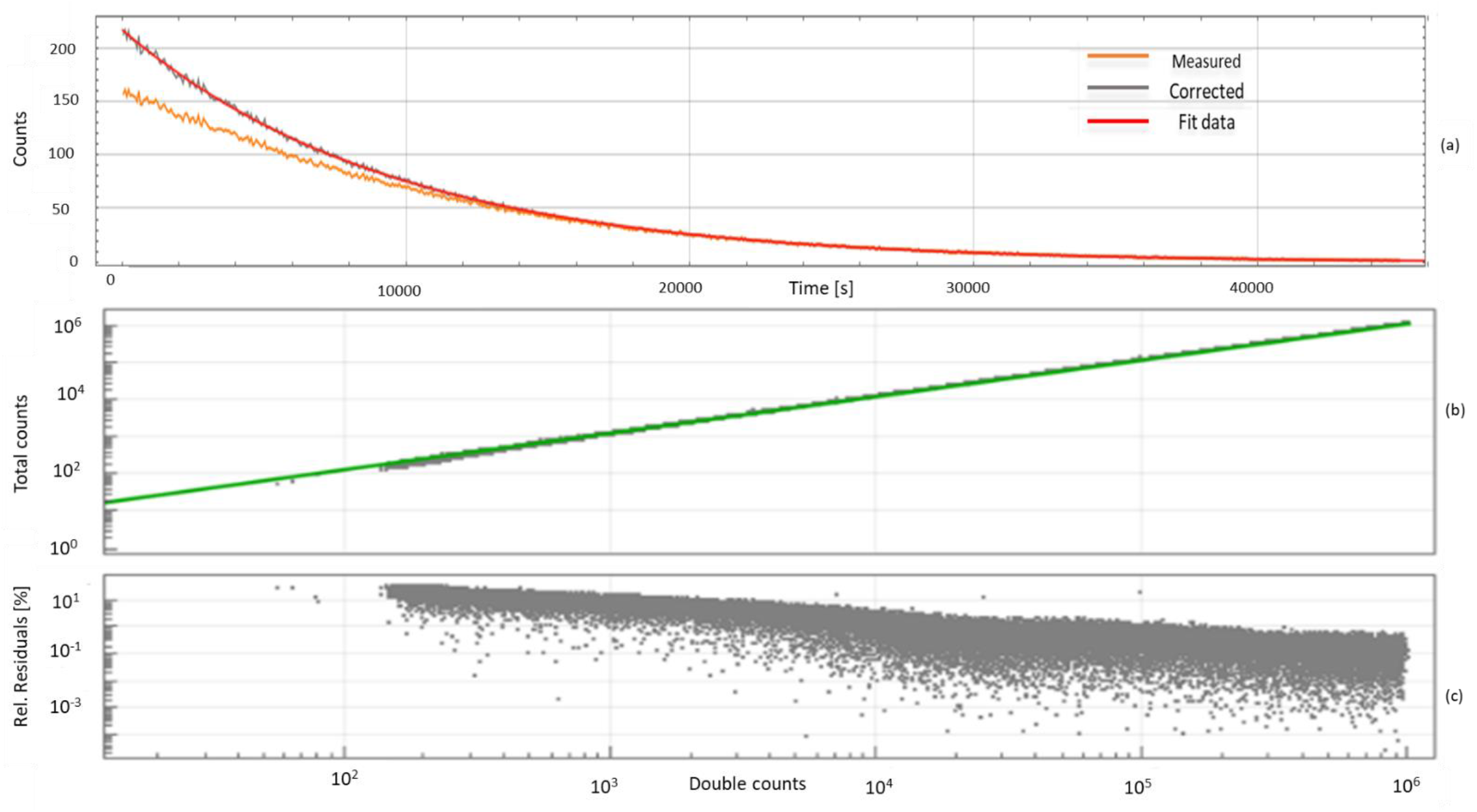
(a) Measured and corrected true coincidences for a single block pair. The red line represents the fit with a single exponential decaying at the rate of ^18^F. (b) The best fit (green line) with an exponential model of the averaged delayed triple and double counts vs the averaged delayed double counts for the decay experiment with ^18^F, the green line is the exponential model. (c) The relative residuals for (b).

**Figure 4.**
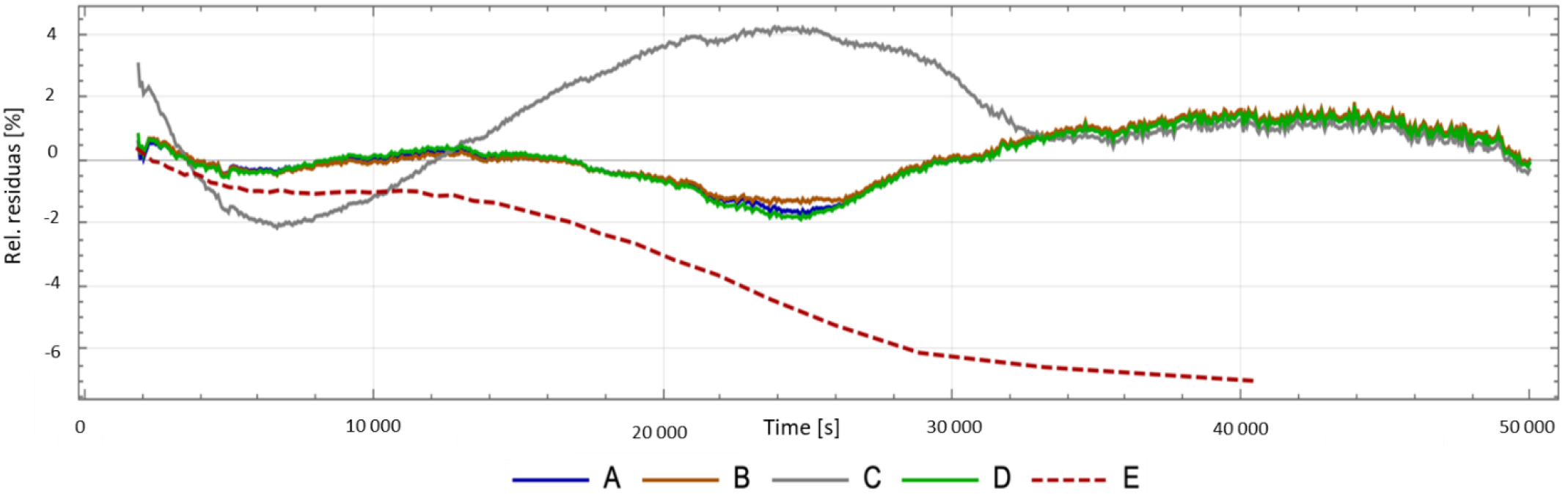
Relative fit residuals of the averaged corrected true coincidences vs time during a phantom experiment with decaying ^18^F and achieved with the presented method of DTC (A) After applying DTC assuming non-paralyzing behavior, individual fit parameters sets, and triple correction, (B) Assuming non-paralyzing behavior, an averaged fit parameter set, and triple correction, (C) Assuming non-paralyzing behavior, individual fit parameter sets, and no triple correction, (D) Assuming paralyzing behavior, individual fit parameter sets, and triple correction. For comparison, the corresponding residuals for the global DTC methods are also shown (E).

**Figure 5.**
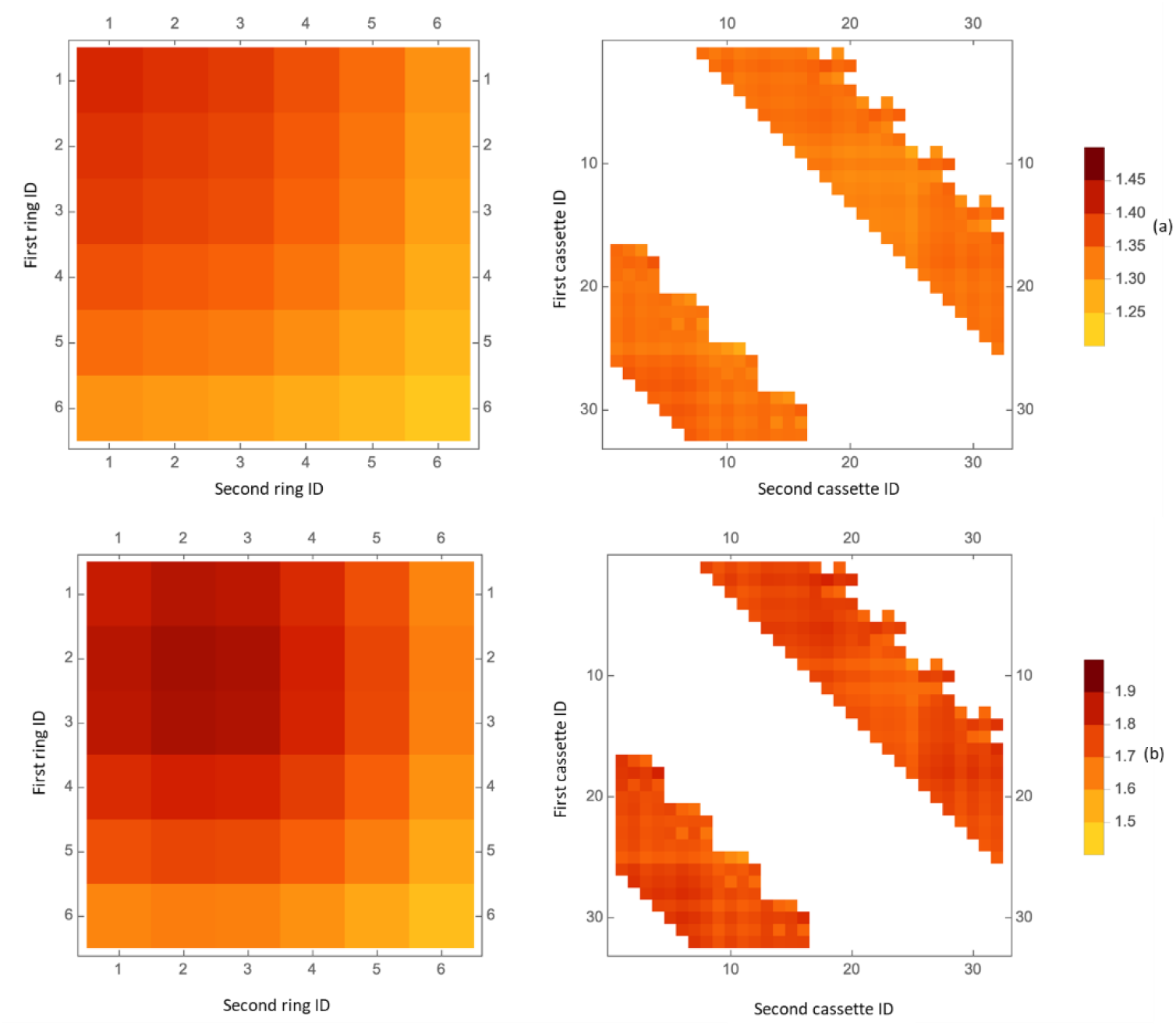
(a) Left: The DTC factors averaged over the first 200 s for different block rings. Right: The DTC factors averaged over the first 200 s for 8 out of 32 detector cassettes, obtained with a phantom measurement inside of the PET FOV and with noticeable outside FOV activity, (b) Left: same as (a), but without any out of FOV activity. White areas in the right plots correspond to ignored lines of response.

**Figure 6.**
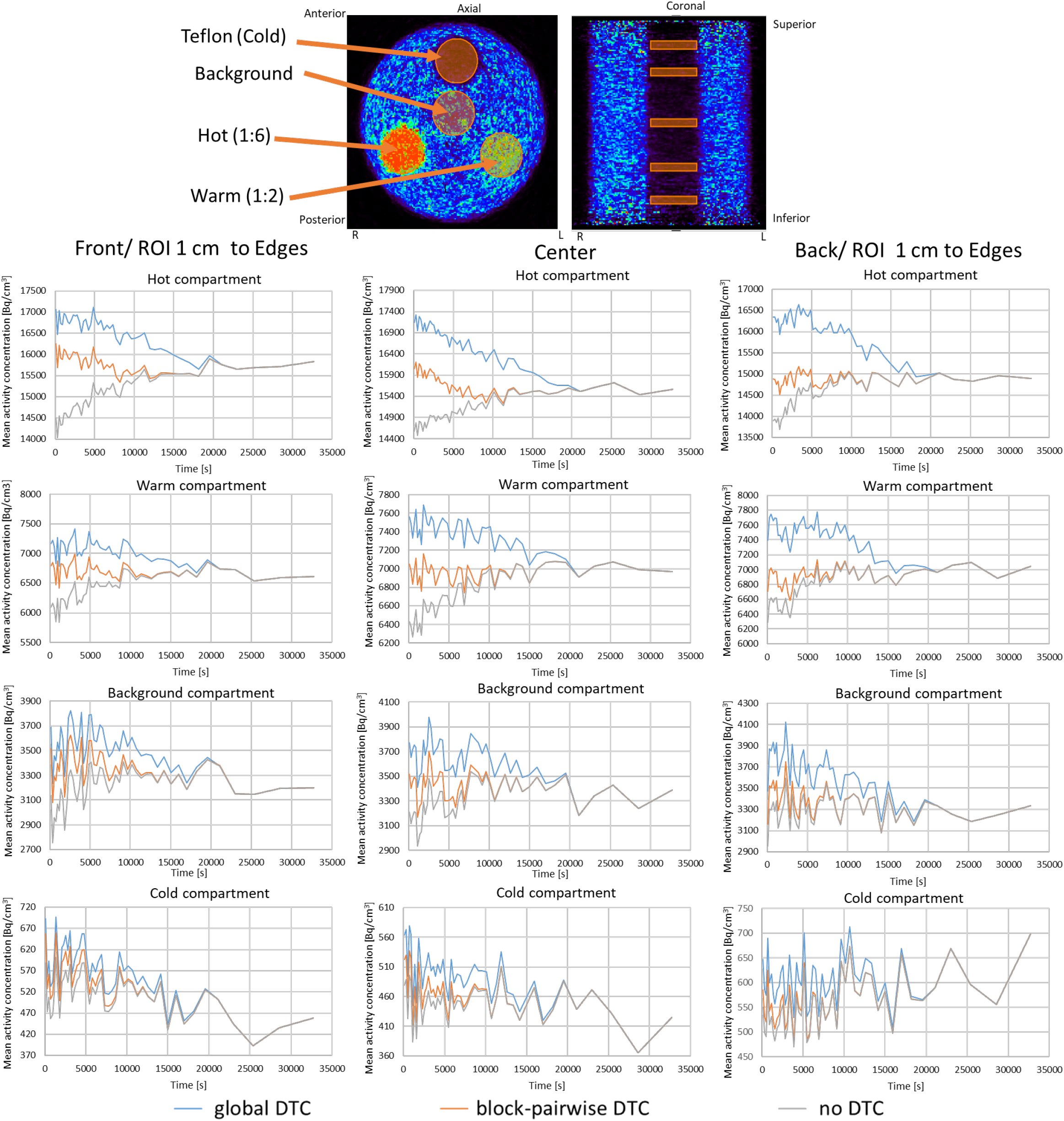
The mean of the activity concentration in Bq/cm^3^ vs. time for the measurement with activity out of FOV with the three compartment phantom for relevant regions.

## 3. Results

The validation of the proposed method showed a very good agreement between the total of expected true counts and the measured and corrected true counts. Figure 3(a) shows the measured and corrected true coincidences for a single block pair together with a fitted model and the corrected true coincidences. An exponential decay with the half-life of ^18^F is assumed. Although only the amplitude of the exponential decay model was adjusted with the least square fit, a very good agreement between the model and measurement was achieved. Figures 3(b) and 3(c) represent the results of the validation of the empirical model for correcting the overestimation of counts due to triple coincidences. Figure 3 (b) presents the overestimation in total prompts due to triple coincidences, i.e. triple coincidences and double coincidences vs. double coincidences, together with a fit under the terms of equation (6). Figure 3(c) shows the relative residual errors for the corresponding fit. Figure 4 shows the fit residuals for fitting the corrected total true coincidence count rates after applying the DTC individually to all individual block pairs and subsequently summing the true counts of all block-pairs. Results for the following four test cases, A to D, are shown.

A: Assuming non-paralyzing behavior and applying the individual sets of the obtained fit parameters to each of the 10944 block pairs.

B: Computing an average set of fit parameters from the individual parameter sets and applying DTC with the average set (assuming non-paralyzing behavior).

C: Assuming non-paralyzing behavior, applying individual parameter sets but omitting triple correction.

D: Assuming paralyzing behavior, applying individual parameter sets, and applying the triple correction.

All relative residuals were obtained by fitting a single exponential decaying at the rate of ^18^F to the global true count rate. In the best case (B), the relative residuals, i.e. the deviation from the ideal behavior, were always larger than ≈ - 1.2% and smaller than ≈ 1.6%. Without triple correction, the maximum deviation from the expected counts was > 2% and < 4%, i.e., substantially larger. Practically no differences were observed between cases (A – B) or (B – D). The remaining part of this work will deal only with DT correction to case (B), i.e., assuming NP behavior, applying an averaged parameter set, and applying triple correction.

Figure 5 shows the DTC factors (averaged for the first 200 s of both phantom measurements with decaying activity. The left image shows the DTC factors averaged over all blocks with the same axial position, thus showing the correction factor depending on the position of the detector ring of the 3T MR BrainPET in which the block resides. The right image shows the DTC factors for the 32 cassettes, with the average over all six blocks in each cassette, thus showing the correction factor depends on the position of the detector’s cassette.The average correction factors were obtained from first phantom measurement (I) with decaying ^18^F inside the FOV (upper figure(a)) and the seocnd measurement (II) with additional decaying activity outside of the FOV (bottom figure(b)). Figure 6 shows the mean activity in Bq/cm^3^ vs. time for the measurement with the three-compartment-phantom and out-of FOV activity. The behavior of the ROI mean and the coefficient of variation are shown for several relevant ROIs. The results without applying any DTC are shown in all plots. The largest differences between the global and the new block-pairwise DTC method were observed in the hot compartment.

Assuming the measured activity concentration for t > 20000 seconds is the actual activity concentration (DT effects can be neglected at the corresponding count rates), global DTC leads to an overestimation of ≈ 6 - 9% in the three regions of the hot compartment, while the overestimation is significantly reduced when using the block-pairwise DTC (only 3% for the reduced when using the block-pairwise DTC (only 3% for the center ROI and minimal and neglectable in the front and back ROI, respectively). For the warm compartment, overestimations with global DTC were reduced to ≈ 6 - 8% in the three regions, while no overcorrection was observed with the block-pairwise DTC method. Overcompensation in the background compartment was of the same order, and lower overcorrections with the block-pairwise DTC method were obtained. Both methods behave very similarly for the cold compartment. Figure 7 shows the COV vs. time for the measurement with the three-compartment-phantom and out-of FOV activity for the same ROIs. The COV is the same for all three DTC methods, i.e. with global DTC, with block-pairwise DTC method, and without DTC. Table 2 shows the results of the differences in the calibration bias by comparing the global DTC method and block-pairwise DTC method with and without activity out of FOV.

**Figure 7.**
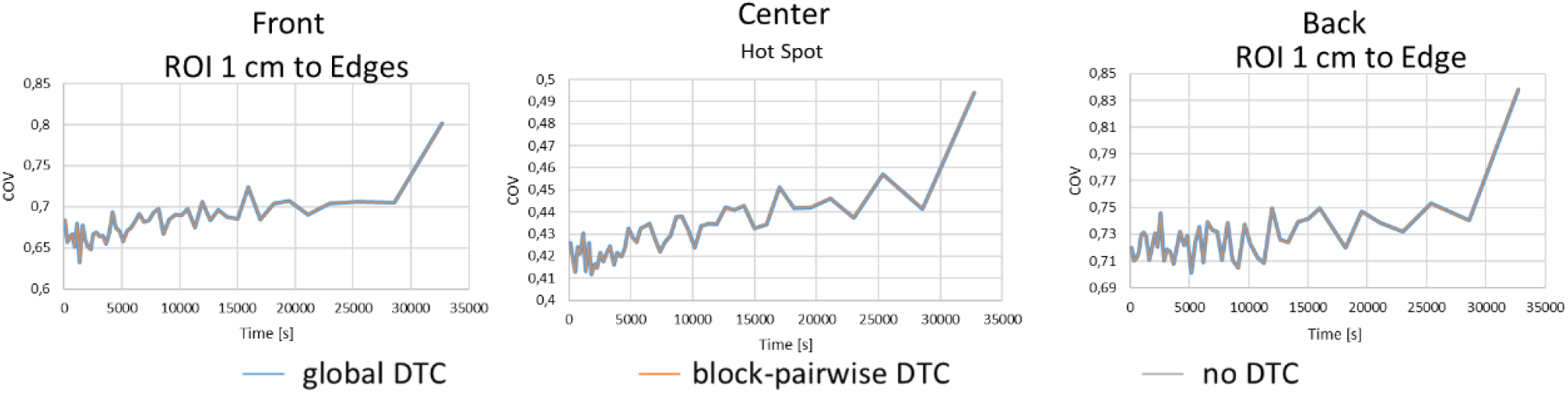
COV vs. time for the measureemnt with the three-compartment-phantom and out-of FOV activity for the same ROIs.

Figure 8(a) shows the TACs in three relevant regions of the human brain, i.e., the cerebellum gray matter, temporal posterior lobes, and ACC from a volunteer measurement with [^11^C]ABP688. Results for global DTC and the new block-pairwise DTC method are shown for comparison. The TACs obtained with both DTC methods show significant differences over the entire scan time. Looking at the BP_ND_ at acquisition times greater than 30 min, i.e. when the steady-state is reached, although the relative differences between block-pairwise and global DTC are small (less than 2%), they still exist and change. A closer look at the TACs over the entire acquisition results in an interesting finding, which is relevant if the distribution volume or the BP_ND_ of a neuroreceptor tracer are determined via kinetic modeling. Figure 8(b) shows ratios between TACs of the ACC and the cerebellum gray matter obtained with global DTC and the corresponding TACs obtained with block-pairwise DTC. This means the overestimation caused by the global DTC, which was also observed in the phantom study, is present here as well. In addition, it is different in ACC and cerebellum gray matter – with a difference of 25% in the case of ACC at early times - and changes over time in a non-linear manner.

**Figure 8.**
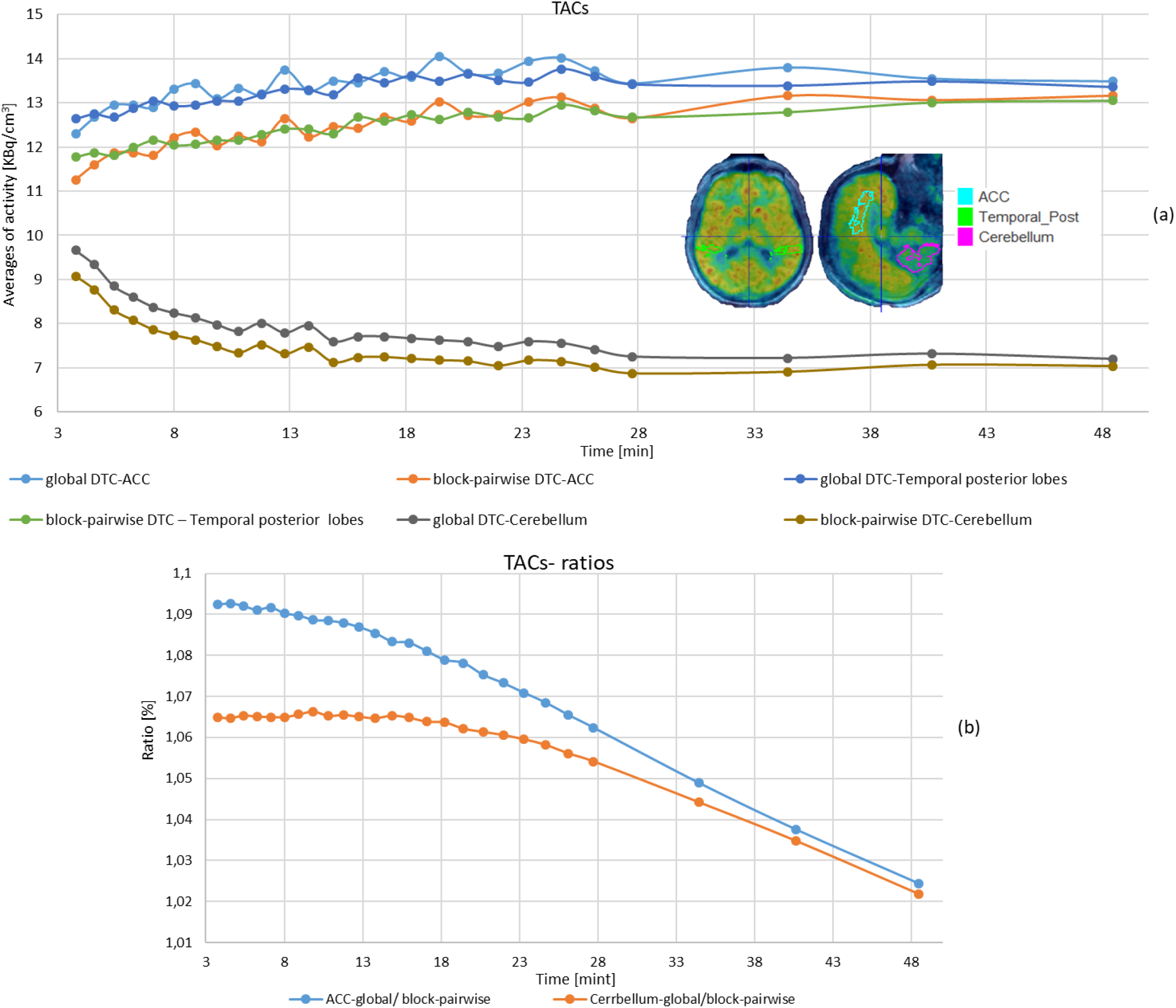
(a) TACs in three relevant regions of the human brain: Cerebellum, Temporal posterior lobe and the ACC from the measurement with [^11^C]-ABP688, for global and block-pairwise DTC, (b) Mean ratios of global to the block-pairwise DTC methods in the ACC to cerebellum, the image reconstructed frames were shortened less than 0.5 lengths to the midpoint since the true counts framing schemes were used.

## 4. Discussion

The presented DTC method is based on the original work from (Yamamoto et al 1986), which aimed to develop a DTC correction scheme independent of the imaged object. In addition to adapting the plane-wise DTC used in (Yamamoto *et al* 1986) to a correction for individual block pairs of the Siemens 3T MR-BrainPET insert, we also introduced measures to account for the correction required for random triple coincidences. Block-pairwise DTC correction was chosen since each individual block detector of the Siemens 3T MR-BrainPET insert with its associated processing circuity (Newport *et al* 2006, Hu *et al* 2011) can be considered as the smallest unit affected by DT. The block detector design of the BrainPET device uses light sharing and a variant of Anger logic for energy and position determination. Consequently, a scintillation event causes DT for the entire block, but it does not cause DT in other blocks of the same ring as in (Grazioso *et al* 2006, Zhang *et al* 2006).

Furthermore, due to the lower total delayed random coincidence in block pairs (10944 in the case of the Siemens 3T MR-BrainPET insert) when compared to the total delayed random counts in a single plane (72 in the case of the Siemens 3T MR-BrainPET insert), special emphasis must be put the on avoiding excessive noise propagation on to the reconstructed image. This was achieved by developing a model (equation (7)) for the observed delayed random events in each pair instead of computing the ratio between measured delayed randoms and extrapolating ideally expected delayed randoms. In this model, DT losses, excess counts due to triple random coincidences, and the natural activity of ^176^Lu (Melcher and Schweitzer 1992, Yamamoto *et al* 2005) were considered.

The effect of correction for triple coincidences can be particularly well observed in figure 4, where the fit residuals obtained by correcting the block-wise true coincidences, their accumulation for the entire system, and the data fitted to a single exponential with a half-life of ^18^F are shown. Without considering excess triple counts, the relative error between the observed DT corrected counts and the ideally expected counts reaches 4%. This error is reduced to a maximum of 1.6% with triples correction. Similar results, i.e., relative deviations of ≈ 1% were achieved in (Yamamoto *et al* 1986). (Vicente *et al* 2013) reported relative deviations of up to 7% with an improved DTC scheme based on the global singles-to-coincidences ratio (SCR) for small-animal PET scanners. (Liu *et al* 2019) described a block-wise SCR DTC method for a long axial FOV PET scanner and reported a maximum bias of 5% in the individual image slices with this method when using two different phantoms (1^st^: homogeneous cylinder, 2^nd^: capillary with scattered). (Liu *et al* 2019), also took potential background radiation into account. Interestingly, there is nearly no difference between DTC corrections for the method presented here when a paralysable and non-paralysable behavior is assumed. Also, using individual fit parameters for all 10944 block pairs does not result in a substantial improvement when compared to using averaged fit parameters. This can be easily understood by the fact that the DTC is dominated by the count rates. The differences for all other fit parameters are most probably only caused by the component tolerances as all 192 block detectors and the associated processing electronics are identical in design. The reason for the remaining systematic deviation from an ideal correction, whose residuals should be purely randomly distributed around 0, is so far unknown and certainly requires further investigation. However, there is an obvious similarity in the systematic modulation of the residuals compared to the case without triple coincidence correction.

Therefore, it can be assumed that the residual systematic deviation from the ideal case is mainly due to an insufficient model for the triple miscount estimation, which was purely empirical in our study, and neglected multiple coincidences. The block-ring-wise and cassette (head)-wise DTC factors shown in figure 5 behaved as expected. With out-of-FOV activity, block pairs inside the first ring required the highest DTC factor, with continuously descending values for pairs in rings two to six. The finding that the maximum DTC factors for the measurement without out-of-FOV activity can be found in ring two and not in ring three or four is due to the fact that the phantom inside the FOV cannot be centered exactly axially because of the Tx/Rx RF head-coil. As expected, no systematic variation in the pairwise DTC factors averaged over all six blocks in a cassette was observed (right figures in figure 5). The evaluation of the accuracy of the DTC methods in inhomogeneous objects, i.e., objects with several compartments containing different activity concentrations, revealed that the global DTC method tends to over-correct the DT losses, especially in the hot compartment.

The lower the activity concentration in the compartment was, the smaller this overcorrection effect was. Against our expectation, the axial position of the ROI did not have a significant impact on the accuracy of the global method. With the new method, only a small overcorrection was observed in the hot compartment at the axially centered ROI. No apparent overcorrection was observed in the warm, background, or cold compartments when using the block-pairwise DTC method. In contrast, the global DTC method produced a significant overcorrection in the warm compartment. The reason for the overcorrection of the block-pairwise method in the hot compartment and the axially centered ROI is not yet fully understood and requires further investigation. We suspect the single scatter simulation (SSS) correction to be responsible for the axial dependency of the block-pairwise method in the hot compartment. This supposition is supported by the residual decrease of the reconstructed activity over time in the cold compartment, even for acquisition times without DT losses, i.e., when the two methods converged to the activity reconstruction without DTC (gray lines). Unfortunately, neither (Yamamoto *et al* 1986) nor (Vicente *et al* 2013, Liu *et al* 2019) evaluated the noise behavior or potential bias in structured phantoms with the presence of out-of-FOV activity. Consequently, a complete comparison of the performance of all mentioned methods was not possible. Results for additional ROIs are given in Supplementary Information (figure 1).The observed differences and the lack of accuracy of the global DTC method have to be taken into account for the regular cross-calibration of the BrainPET scanner, as the overcorrections of the global method based on CFD counts may lead to an overestimation of the calibration factor, which has to be especially considered when comparing images that differ in their DTC method and for high activity levels where DTC is still required. Although the relative deviations between calibration values with and without out-of-FOV activity are smaller for the block-pairwise DTC method compared to the global method (see table 2), there is still a residual inaccuracy of the block-pairwise method, which we also attribute to the inaccuracies of the SSS correction.

The observed inaccuracies of the global DTC method can lead to relevant quantitation errors in human studies, as demonstrated in figure 8. The [^11^C]-ABP688 study, the data from which were utilized here, was performed with a bolus-infusion protocol, meaning that the BP_ND_ was determined after 30 min. Although, in this case, the effect of the global DTC compared to the block-pairwise DTC was rather small, there was a 2% difference in the BP_ND_. This difference can potentially mask the effects of cognitive challenges as described in (Régio Brambilla *et al* 2022), as it decreases over time and alters for different brain regions. Despite not being relevant for the present bolus-infusion protocol investigation, the early part of the TACs allow an important conclusion for dynamic studies using a bolus injection protocol and kinetic modeling to be drawn, which requires evaluation of the entire TAC to derive the BP_ND_ outcome. Figure 8(b) shows that the overestimation of the early TACs caused by the global DTC is much greater for the ACC than for the cerebellum. Furthermore, this overestimation is not constant and changes, i.e., decreases, over time. This means that the prerequisite for the kinetic modeling of dynamic studies concerning spatial and temporal invariance is violated. In a typical dynamic neuroreceptor study with a bolus injection protocol, the difference between (early) high counts and (late) low counts in the course of the acquisition can be expected to be even greater than in the present [^11^C]-ABP688 study.

Finally, the observations and considerations presented in this work may also be relevant for DTC in other PET systems. Although, based on the geometry and the positioning of the object, we would expect a less significant impact on the insufficiencies of global DTC, the effects should be studied in detail in an independent research work. Despite this, the presented results are equally relevant for other dedicated brain PET devices which have been developed in recent years or are currently under development, e.g. (Gonzalez *et al* 2018, Jung *et al* 2015, Nishikido *et al* 2017, Won *et al* 2021, Del Guerra *et al* 2018, Teimoorisichani and Goertzen 2018, Moliner *et al* 2019, Ahnen et *al* 2020, Catana 2019, Lerche *et al* 2020, Carson *et al* 2021).

## 5. Conclusions

This work presents a more accurate DTC method for a 3T MR BrainPET insert. The method is based on the estimation of the DTC factor from the delayed random coincidences between individual block pairs and includes a correction for detected random triple coincidences and the ^176^Lu background. The method has been evaluated with measurements using phantoms with compartments filled with different activity concentrations and with out-of-FOV activity. The quantification bias was significantly smaller in all evaluated compartments and ROIs when using the new method. We also demonstrated that the dependency of the calibration factor on out-of-FOV activity could be reduced compared to the global DTC method. The increased quantitation accuracy, which is achieved with the improved DTC method, is potentially highly relevant for dynamic neuroreceptor studies that aim to quantify tissue parameters such as the distribution volume and the BP_ND_. When comparing the global DTC method to the newly developed method, we observed relative differences in the ACC and cerebellum gray matter - with a difference of 25% in the case of ACC at early times and non-linear changes over time. These differences could lead to erroneous results in kinetic modeling, e.g., the masking of the effects that cognitive tasks have on the TACs in the ROI and the reference region.

## Data Availability

All data produced in the present work are contained in the manuscript

## Acknowledgment

Open access publication funded by the Deutsche Forschungsgemeinschaft (DFG, German Research Foundation) – 491111487. This work was supported by the German Federal Ministry of Education and Research (BMBF grant 01DH16027) within the framework of the Palestinian-German Science Bridge project.

