## Supplementary material for "A Detector Block-Pairwise Dead Time Correction Method for Improved Quantitation with a Dedicated BrainPET Scanner": Supplentental Figure1Figure2

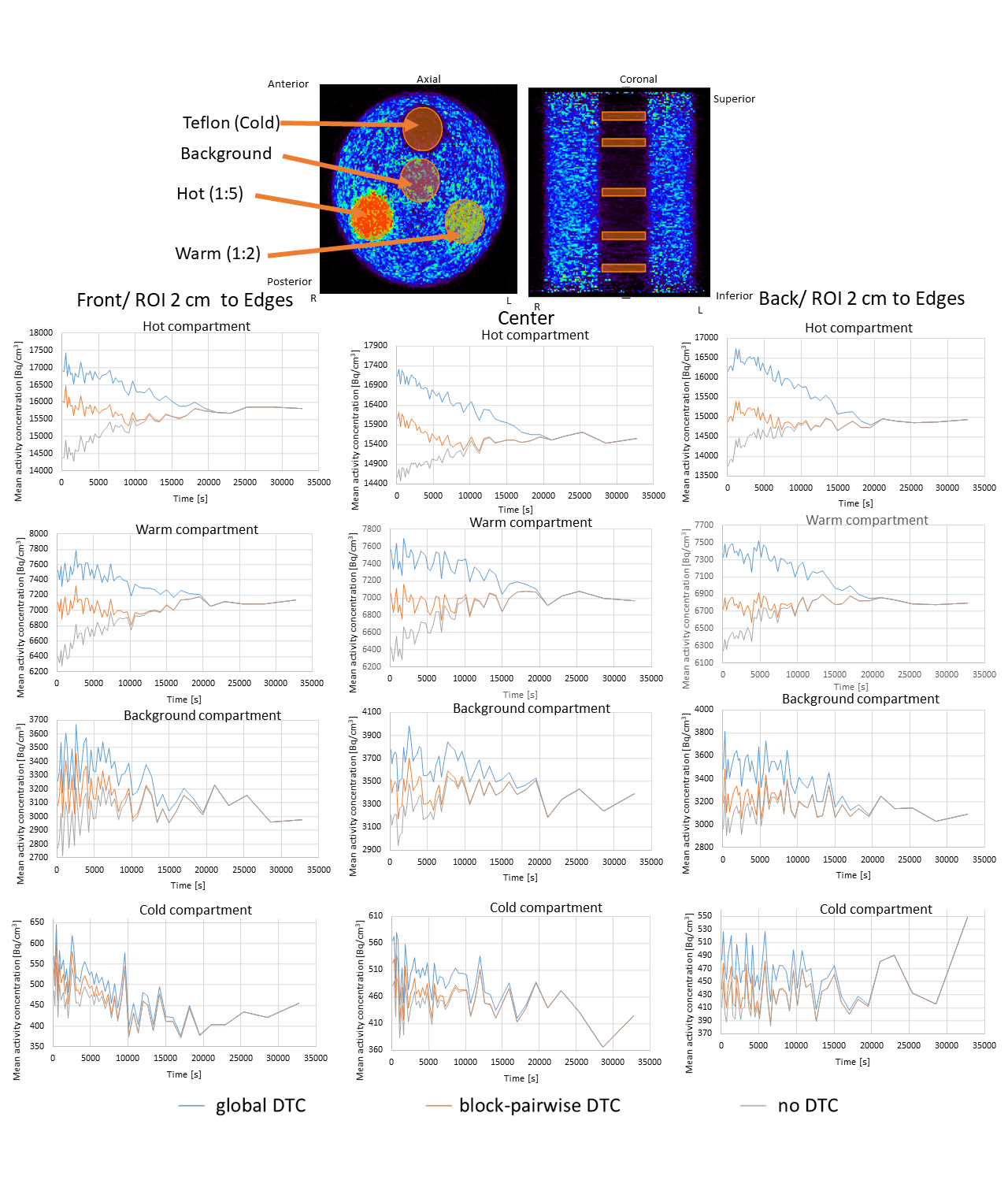
**Figure.1** The mean of the activity concentration in Bq/cm3 vs. time for the measurement with activity outside of FOV with the three-compartment phantom for relevant regions.

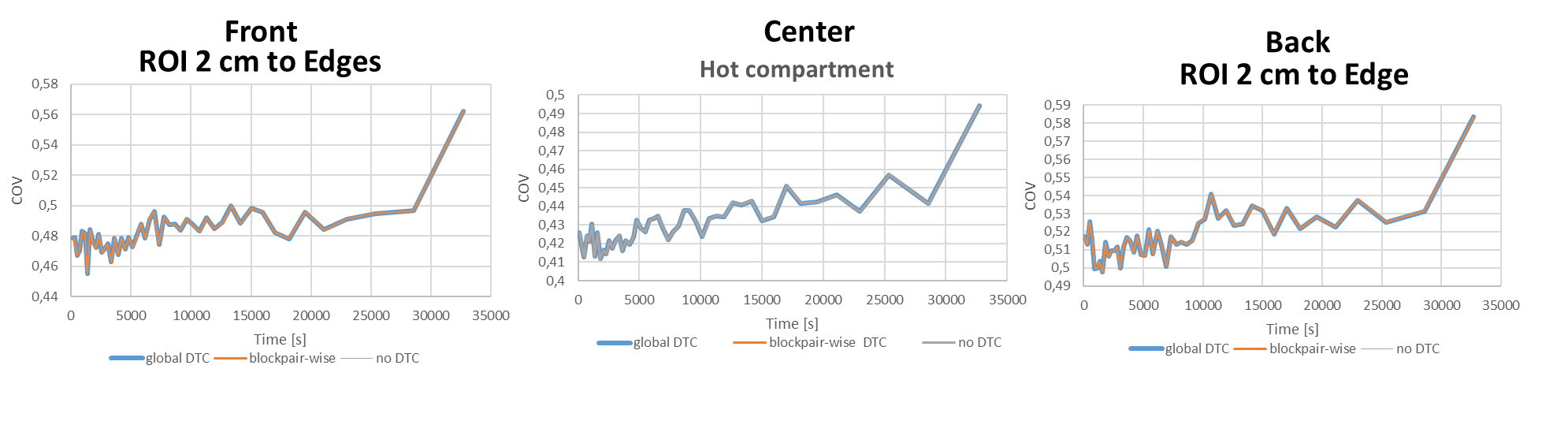

**Figure. 2** COV vs. time for the measurement with the three-compartment phantom and out-of FOV activity for the same ROIs.
